# Exploring the Connection Between Pet Attachment and Owner Mental Health: The Roles of Owner-Pet Compatibility, Perceived Pet Welfare, and Behavioral Issues

**DOI:** 10.1101/2024.11.20.24317636

**Authors:** Roxanne D. Hawkins, Annalyse Ellis, Charlotte Robinson

## Abstract

Research exploring the connection between pet ownership and mental health has expanded substantially in recent years, yet scientific evidence remains inconclusive. Existing studies have oversimplified this relationship by focusing primarily on pet ownership itself, without accounting for crucial factors such as species of the pet, or important relationship dynamics such as owner-pet attachment orientations. This study sought to investigate whether the relationship between owner-pet attachment and owner mental health could be better understood through the lens of owner-perceived pet compatibility, perceived pet welfare, and pet behavioral issues. These aspects, often overlooked in previous research, are believed to play crucial roles in shaping owner-pet relationships and owner mental wellbeing. This study surveyed emerging adults (ages 18-26) from the UK (N=600) with anxiety and/or low mood who owned dogs and/or cats. A large portion of the sample met clinical criteria for Generalized Anxiety Disorder or Major Depressive Disorder. Our findings revealed that dog owners exhibited more secure pet attachments than cat owners. Attachment notably influenced mental health whereby anxious attachment was linked to poorer mental health among dog owners, while avoidant attachment was associated with better mental health in both dog and cat owners. Insecure attachment related to poorer pet quality of life, increased reports of pet behavioral problems, and poorer owner-pet compatibility, regardless of pet species. Additionally, poorer welfare and more behavioural problems were associated with poorer mental health for dog owners; these findings were not replicated for cat owners. Notably, a dog’s mental state (such as appearing depressed), as well as fear and anxiety in dogs, mediated the relationship between owner-pet attachment and owner mental health. Owner-dog compatibility, particularly in the affection domain, influenced owner anxiety, positively mediating the relationship between anxious attachment and poorer mental health, while negatively mediating the relationship between avoidant attachment and better mental health. These findings suggest that a simplistic view of pet ownership fails to capture the complexity of the factors that shape the mental health of pet owners and underscores the need to consider important owner-pet factors to fully understand how the human-pet relationship can impact the wellbeing of both people and their pets.

## Introduction

Emerging adulthood, which approximately comprises ages 18 to 26 [1], is a distinct and transitional life stage bridging the adolescent and adult life stages [2, 3]. During emerging adulthood, individuals often make important decisions related to several consequential areas of life such as education, work and career, identity, love, and family [2] and undergo continued rapid brain development [4]. Due to the transitional element of this period of life, emerging adults often experience mental wellbeing concerns such as lower life satisfaction [5], increased stress [6], loneliness [7], and increased vulnerability to psychological disorders [8, 9]. Emerging adulthood is considered a peak age for the onset of mental health difficulties, with approximately 75% of mental health disorders being diagnosed between the ages of 18 and 25 years, particularly anxiety and depression [10]. During this tumultuous time, social support and secure attachments can serve as protective factors related to wellbeing [11–13]. One such source of support and protective factor during this life period may be companion animals, hereby referred to as ‘pets’.

Pets are often acquired for companionship [14], are viewed as a provider of social support by many owners [15], and often play an important role in the daily management of mental health problems [16]. In emerging adulthood, high importance is often placed onto relationships and meaningful interactions with pets; pets can be self-management tools for emotional health, helping to reduce loneliness and anxiety, and promoting coping and resilience, particularly for young adults facing hardships and adversity [17–19]. Within emerging adulthood, existing literature has indicated that the human-pet relationship can have a positive association with wellbeing. For example, affection from a companion animal has been associated with better psychosocial functioning [19], and for LGBTI+ young adults, pets may promote feelings of belongingness, positive regard, and emotional support [17]. However, some literature also highlights a negative association between pet ownership and wellbeing, such as pet ownership being reported as a source of stress and caregiving burden for emerging adults [17]. For university students, people substituting-related pet attachment, more frequent co-sleeping with pets, and spending more time with pets prior to leaving home for university was associated with increased separation anxiety while away from home [20]. The mixed outcomes observed is not uncommon within the human-animal field in general, and despite an increase in research interest, findings relating to the “pet effect,” or the well-being benefits derived from pet ownership, have been highly inconsistent across the lifespan [21, 22]. Such inconsistencies in findings may possibly be explained by the lack of consideration for the complex interplay of individual factors within human-pet relations that can confer both risk and benefit for mental health [23].

Pets may exacerbate pre-existing mental health difficulties through increased worry and stress, particularly in cases whereby individuals feel they are not meeting the needs and expectations of their pet (or vice versa), do not feel satisfied and fulfilled by a relationship, or whereby the pet has health or behavioural issues that cause concern and distress for owners [24–27]. A relationship with a pet may therefore be either particularly beneficial or stressful, depending on other variables related to pet ownership, such as the nature and markers of the quality of the human-pet relationship such as attachment [28–30], perceived compatibility [31], and individual differences in pet-related factors such as health and behaviour [32]. Moreover, pet species differences may be an important factor. For example, more positive outcomes in relation to wellbeing have been observed for dog owners, although it should be noted that there is limited research into the wellbeing outcomes of cat owners [33, 34]. Human-dog attachments are, however, bi-directional and there is reciprocity within the relationship, which is important for wellbeing [35, 36], yet less is known regarding the nature of human-cat relationships, which may be more variable, and where different attachment features may be observed [37, 38]. The present research seeks to better understand the interactions among these variables and their associations with well-being outcomes for the emerging adult population.

### Relationship markers: Attachment

Attachment theory was initially developed to describe the relationship between parent and child, wherein relationships may be marked by security or insecurity [39]. Attachment security is the extent to which an individual feels assured that their emotional needs will be reliably met by others [40]. Attachment insecurity is a two-dimensional construct, consisting of both attachment anxiety and attachment avoidance. In human-human relationships, attachment anxiety describes the fear or anxiety that others will not be responsive to emotional needs, while attachment avoidance describes the emotional distance caused by worry regarding others’ intentions [40]. Similar features of attachment insecurity have been observed within human-pet relationships [41, 42]. Attachment has been extensively researched within human-human relationships but has only more recently been applied to the exploration of the inner dynamics of the human-pet relationship. There are salient challenges related to the measurement of this construct within the human-pet relationship. Although there are many studies claiming to have measured human-pet attachment, many of these studies may be measuring the strength of the human-animal bond, rather than a two-dimensional model of attachment. A recent systematic review related to human-pet attachment and mental health found that, of studies measuring both attachment and depression, only seven of the forty included studies operationalized attachment in a way that aligned with psychological attachment theory [43]. More research is therefore needed to further explore the association between attachment orientation within human-pet relationships and human wellbeing outcomes.

Research on the long-term developmental and psychological benefits of secure attachments within human-human relationships is well-established, yet little is known regarding attachment within human-pet relationships, despite these animals being considered as close family members by many owners and being ranked high in attachment hierarchies [15]. Studies that have investigated the human-pet attachment bond and owner wellbeing have indicated that there may actually be heightened emotional vulnerability in those who are strongly attached to their pet, with attachment scores being positively correlated with psychological distress through increased loneliness, depression, anxiety, somatoform symptoms, and mental health burden, along with a fear of rejection and being unloved [44–46]. However, these studies only examined the human-pet attachment bond, rather than attachment orientation (e.g., secure/insecure). The few studies that have considered attachment orientation and owner wellbeing within the pet domain indicate that feelings of security within pet attachments are associated with lower psychological distress and psychopathology and better psychological health, such as fewer depressive symptoms [30, 47]. Anxiety within pet attachments has been previously associated with poorer owner mental wellbeing, whereas avoidance in pet attachment was not found to be related to owner mental wellbeing [30, 48]. Given the proposed similarities observed within human-human and human-pet attachment orientations, particularly in the context of pets providing a safe and secure base and fulfilling the role of an attachment figure [28, 41, 42], we can hypothesise that similar psychological benefits may be observed within secure human-pet attachments. Similarly to human-human relationships, human-pet relationships are both rich and complex, and there could be important mechanisms impacting upon the quality of these relationships and therefore mediating the association between attachment and wellbeing, including owner-pet compatibility, perceived pet welfare, and pet behaviour problems.

### Relationship markers: Compatibility

One important relationship marker, human-pet compatibility, has received little attention in the existing literature, but may play an important role in the relationship between attachment and the derivation of well-being benefits from the human-pet relationship. In human-human relationships, compatibility is a notable component of successful relationships, contributing to higher relationship quality and relationship satisfaction [49–51], which robustly predicts well-being [52]. Similarity in physical and psychological characteristics can influence partner choice and can help maintain a successful relationship; there is also consistent evidence for the ‘similarity-attraction hypothesis’ (i.e. higher attraction between two individuals when there is high similarity across multiple domains) within the human-human relationship field [53]. This hypothesis may also apply to human-pet relationships [54]. High compatibility, or ‘matches’ between owners and pets could promote positive functioning human-pet dyads, strengthen bonds, promote relationship satisfaction, and prevent pet relinquishment [31]. Owner-pet compatibility relates to better owner mental health [55, 56], and pet owners who feel that they have similar characteristics to their companion animals also report better well-being [57]. Additionally, feelings of similarity and reciprocity in terms of need fulfilment within a relationship (whether actual or perceived), enjoyment from shared activities, and pet characteristics that compliment an owners’ needs and lifestyles (e.g., activity preferences), are important to flourishing relationships, and could therefore influence the psychological benefits derived from pet ownership [27, 35, 58, 59]. However, research into owner-pet compatibility has largely focused on matches or mismatches between owner and pet personality factors (e.g., [54, 60]), thus focusing on the psychological dimension of compatibility; yet compatibility also contains physical, emotional, social, and behavioural dimensions. Examining a wider range of compatibility dimensions is important to identify which are most important for human-pet attachment and owner mental health and has implications for strategies to promote more successful and functional human-pet dyads. Existing research has also largely focused on human-dog dyads, with human-cat relationships being overlooked. In sum, there is a tenuous relationship between human-pet compatibility and owner well-being that needs further investigation in relation to both dog and cat ownership.

### Pet-related factors: Pet behaviour and welfare

Past research indicates that a positive pet attachment bond is linked to increased caregiving behaviours, which may improve welfare outcomes for pets [61, 62]. The present study aims to expand upon this research, exploring the role of attachment anxiety and avoidance within human-pet relationships. It is possible that individuals higher in attachment anxiety may feel more positive emotion when engaging in caregiving tasks, and subsequently provide more care to their pets [63–65]. In contrast, individuals higher in attachment avoidance find less meaning and feel lower levels of positive emotion when engaging in caregiving tasks [64], and subsequently may be less caring towards their pets [66, 67]. Perceived pet welfare may also mediate the relationship between attachment and pet owner wellbeing. In human-human relationship research, a child’s wellbeing can have an impact upon the wellbeing of their parents [68], and a romantic partner’s wellbeing can impact upon one’s own health and wellbeing [69]; thus, it is possible that these findings may extend to human-pet dyads. Poor pet welfare, preoccupation with potential pet loss, and anticipatory grief may increase stress and anxiety in owners, exacerbating owner mental wellbeing concerns [25]. There are also emotional and psychological effects of caring for a sick pet which can increase psychosocial distress and clinically meaningful caregiver burden, which in turn can increase symptoms of depression and anxiety [70, 71]. Along with concerns regarding a pet’s welfare, perceived behavioural problems in pets can be seriously disruptive to the human-pet relationship, resulting in worse mental health outcomes for the pet owner [24, 72], as well as pet relinquishment and abandonment [73, 74]. Pet behaviour problems are associated with a range of negative emotions experienced by pet owners, such as anger, stress, and sadness [72], decreased life satisfaction and wellbeing [71], and mental health concerns such as depression and anxiety [24]. However, it should be noted that ‘problem’ behaviours reported by owners may be natural instinctive behaviour of the pet species (e.g., digging), and so it may be the owner’s *perception* that these behaviours are undesirable that could impact upon owner stress and wellbeing; there are also individual differences in which behaviours are viewed as undesirable (e.g., jumping may be viewed positively by some owners) [71]. Moreover, an owner’s attachment may influence pet behaviour through differences in owner behavioural strategies during challenging situations [75]. For example, owner attachment insecurity has been associated with increased dog aggression and separation related disorders [76, 77]. Given the potential impact of owner-perceived pet welfare and behaviour on owner wellbeing, these under looked variables were important to consider when examining the relationship between human-pet attachment, owner wellbeing in the current investigation.

### The present research

Despite the existing literature related to pet attachment and owner wellbeing, there are still consequential gaps in the human-animal interaction field’s understanding of markers of relationship quality that could be impacting upon owner wellbeing. This study seeks to address those gaps by answering the following research questions: 1) Are there differences between dog and cat owners on measures of pet attachment and mental health?; 2) Does insecurity in pet attachment relationships relate to owner mental health symptom severity?; and 3) Does perceived pet welfare, pet behavioural problems, and perceived owner-pet compatibility explain the relationship between insecure pet attachment and owner mental health symptom severity?

## Method

### Design and Procedure

Participants were recruited through Prolific, an online recruitment service, a highly efficient recruitment method to ensure high-quality data with a quick turnaround. A balanced sample was requested for an equal distribution of gender. Participants were screened through filters, and the inclusion criteria included: 1) aged 18-26 years, 2) nationality and area of residence was the United Kingdom, 3) first and fluent language is English, 4) have a dog and/or cat, 5) identified as having difficulties with anxiety and/or depression/mood. Exclusion criteria included: 1) incomplete survey submissions, 2) survey completion times that were three standard deviations below the mean, and 3) failing more than one attention check; in line with Prolific guidance and recommendations.

An online survey was hosted on the Qualtrics platform and eligible participants were directed to the survey through their Prolific dashboard. Participants first read an online participant information sheet and provided informed consent prior to viewing the survey questions. Once surveys were completed, participants were de-briefed and re-directed to Prolific through linking Qualtrics to Prolific with URL re-directs. Given the sensitive nature of the topic, participants were provided with mental health resources both prior to the survey and during the de-brief. The survey took a median length of 14 minutes. Participants received payment through Prolific using the standard tariff (£3 per 30 mins) which follows ethical pay practices and is in line with the minimum reward per hour reward policy. Payments were only made to anonymous participants recruited via Prolific.

### Participants

Priori power analysis: a minimum sample size of *N*=68 was required to achieve 80% power in detecting a medium effect size based on alpha of .05 for each mediational analysis. A total of 656 participants responded to the survey, from this, *n*=50 was returned (i.e. did not meet inclusion or exclusion criteria), and *n*=6 timed out of the survey. The final sample included *N*=600 with *n*=341 dog owners, and *n*=259 cat owners.

Ages ranged from 18-26 years (M=22.57, SD=1.90). Some participants had multiple cats (range 1-8, M=2, SD=1) and/or multiple dogs (range 1-6, M=2, SD=1) but were asked to answer the study questions based on the pet they currently lived with, felt the most attached to, or had owned the longest. Length of pet ownership ranged from under 6 months to over 10 years, and pet age ranged from under 1 year to over 10 years. Most participants lived with their pet full-time at the time of the study (*n*=505), but due to the participant age group, some participants lived with their pet part-time (*n*=83) or did not live with their pet at the time of the study (*n*=12) (e.g., living away from home at university). We aimed to recruit young adults who were having difficulties with their mental health (low mood/depression and/or anxiety); a mental health diagnosis was not required due to low help seeking numbers previously found within this population [78]. Participant demographic information can be found in Table 1.

**Table 1.**
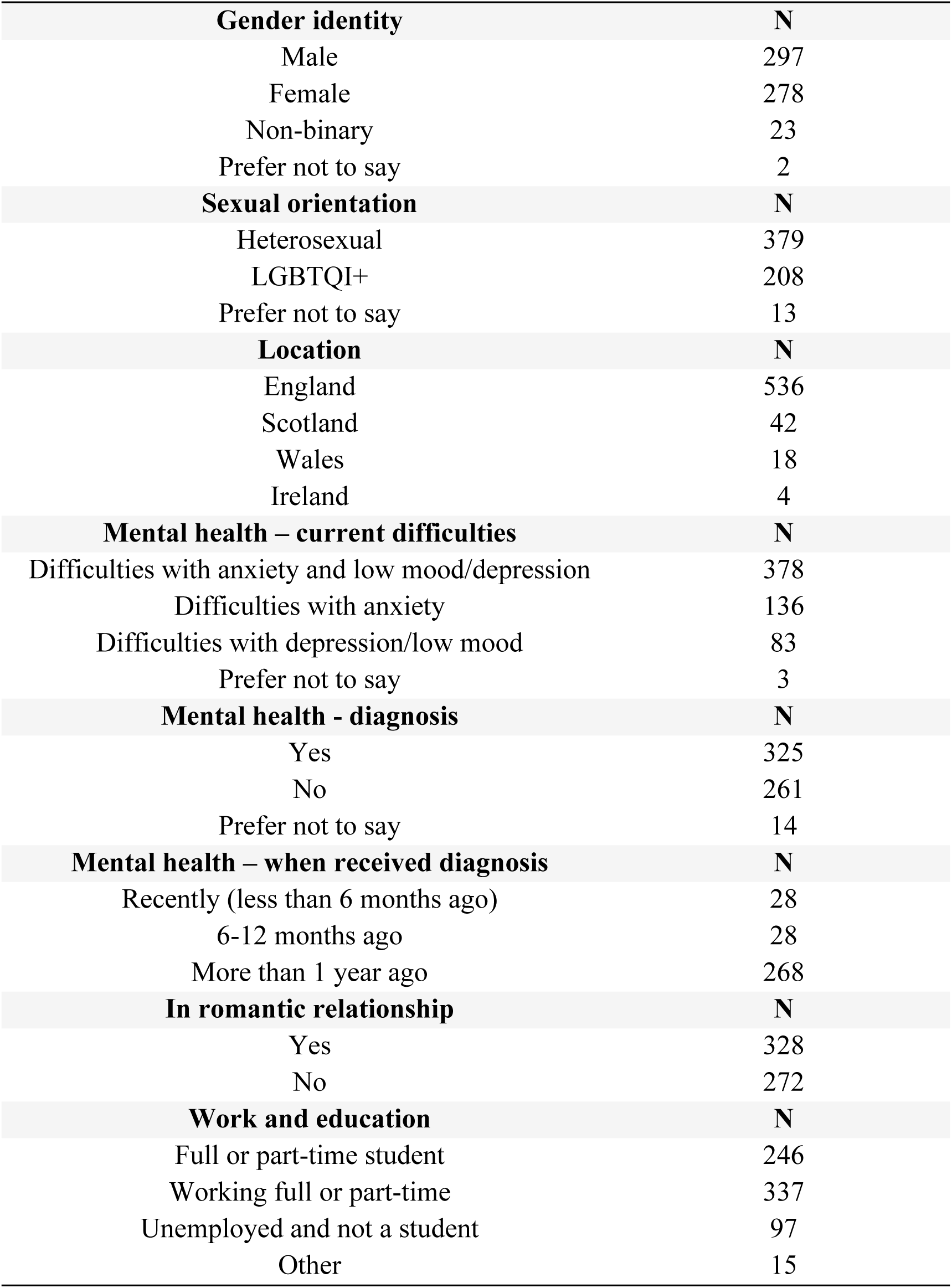
Participant demographics.

| **Gender identity** | **N** |
| --- | --- |
| Male | 297 |
| Female | 278 |
| Non-binary | 23 |
| Prefer not to say | 2 |
| **Sexual orientation** | **N** |
| Heterosexual | 379 |
| LGBTQI+ | 208 |
| Prefer not to say | 13 |
| **Location** | **N** |
| England | 536 |
| Scotland | 42 |
| Wales | 18 |
| Ireland | 4 |
| **Mental health – current difficulties** | **N** |
| Difficulties with anxiety and low mood/depression | 378 |
| Difficulties with anxiety | 136 |
| Difficulties with depression/low mood | 83 |
| Prefer not to say | 3 |
| **Mental health - diagnosis** | **N** |
| Yes | 325 |
| No | 261 |
| Prefer not to say | 14 |
| **Mental health – when received diagnosis** | **N** |
| Recently (less than 6 months ago) | 28 |
| 6-12 months ago | 28 |
| More than 1 year ago | 268 |
| **In romantic relationship** | **N** |
| Yes | 328 |
| No | 272 |
| **Work and education** | **N** |
| Full or part-time student | 246 |
| Working full or part-time | 337 |
| Unemployed and not a student | 97 |
| Other | 15 |

### Survey measures

Following socio-demographic and pet owning questions, a range of standardized and validated measures were presented, investigating owner-pet attachment, owner-pet compatibility, anxiety, depression, pet behavioural problems, and pet welfare. Some measures were specific to dog or cat ownership.

#### Owner-pet attachment

A limitation of other known pet attachment measures is that they do not align with psychological attachment theory and therefore may not be capturing an attachment relationship. The Pet Anxiety and Avoidance measure [28] was chosen for this study because it was developed from a well-utilised measure for human attachment which taps into the two-dimensional model of attachment in adults, examining both anxiety and avoidant attachment orientations (ECR-R) [79], (RQ) [80]. The measure includes a 16-item scale, each scored on Likert scale from 1-7 (‘Strongly Disagree’ to ‘Strongly Agree’). The measure has two subscales: the Pet Avoidance Scale, and the Pet Anxiety Scale, with 8-items each. Scores are continuous, and mean scores are calculated. The measure has been shown to be reliable in previous studies [28]. Cronbach’s alphas for this study: Total (α = .83), Anxiety (α = .78), Avoidance (α = .82).

#### Owner-pet compatibility

The Human-Dog Behavioural and Emotional Compatibility (HDBEC) measure is a new measure developed for this study, adapted from González-Ramírez [81]. This measure consists of 20-items to evaluate human preferences (“I enjoy / would enjoy…”), and 20-items to evaluate dog preferences for the same activities (“Your dog enjoys…”), categorised into five domains of compatibility with 4-items each: Physical (e.g., “To exercise with my dog, e.g., running, hiking, walking, swimming”), Social (e.g., “For me and my dog to meet and interact with new people/strangers”), Affection (e.g., “To stroke, pet, and touch my dog”), Closeness (e.g., “To have my dog with me when I relax, e.g. watch tv, read a book”), and Other (e.g., “To take pictures/videos of my dog”). Participants are asked to “Please choose the response that best fits you and your dog” and each item is rated on a 4-point Likert scale: 0 ‘Totally Disagree’ to 3 ‘Totally Agree’. Scores are compared for owner preference and dog preference for each item, a score of 2 is given for an exact match, and a score of 0 for no match. A total compatibility score is calculated for each domain (range 0-8) and across domains (range 0-40). Higher scores indicate higher compatibility. Cronbach’s alphas: Total (α = .90), Physical (α = .78), Social (α = .81), Affection (α = .79), Closeness (α = .81), Other (α = .74). The measure is openly available for use [https://osf.io/s5ejy/].

The Human-Cat Behavioural and Emotional Compatibility (HCBEC) measure is a new measure developed for this study, adapted from the HDBEC. The measure structure is the same as the dog measure, also consisting of 20-items for both human and cat preferences, categorised into the same five domains of compatibility: Physical (e.g., “To play with my cat, e.g., games, toys, ball, hide and seek”), Social (e.g., “For my cat to initiate social interactions with me, e.g., nudges me, paws at me, is vocal)”, Affection (e.g., “To stroke, pet, and touch my cat”), Closeness (e.g., “When my cat stays close / follows me”), and Other (e.g., “To take pictures/videos of my cat”). This measure is scored and coded the same way as the dog measure. Cronbach’s alphas: Total (α = .90), Physical (α = .66), Social (α = .66), Affection (α = .80), Closeness (α = .84), Other (α = .75). The measure is openly available for use [https://osf.io/s5ejy/].

#### Pet welfare

A direct quality of life (QOL) assessment was included that asked participants “How would you rate your pet’s current quality of life?” rated on a scale of 1 “Very poor” to 10 “Excellent”. Total scores are calculated.

For dogs, the Canine Health-Related Quality of Life Survey (CHQLS-15) [82] was included that is comprised of 15-items that assess owner perceived dog quality of life through four domains: Happiness (e.g., “My pet enjoys life”), Physical functioning (e.g., “My pet moves normally”), Hygiene (e.g., “My pet keeps him/herself clean”), and Mental status (e.g., “My pet seems dull or depressed, not alert”). Participants are asked to think about the past four weeks when rating each item. Each item is scored on a 5-point Likert scale from 0 “Never / Strongly disagree” to 4 “Always / Strongly agree”. Mean scores are calculated for each domain as well as across domains for a total HRQoL score (range 0-4). Cronbach’s alphas: Total (α = .83), Happiness (α = .70), Physical functioning (α = .70), Hygiene (α = .60), Mental status (α = .54).

For cats, the Feline Health-Related Quality of Life (FHQLS) measure first asks participants “Thinking about the past 4 weeks… the general health of my cat has been..?” which they rate on a 5-point Likert scale from 1 “Poor” to 5 “Excellent”. Participants then score a further 21-items (e.g., “My cat has yowled in distress”) on a 5-point Likert scale from 0 “Not at all / Strongly disagree” to 4 “A great deal / Strongly agree”. Negatively worded items are reverse coded and then total scores are calculated; higher scores indicating higher welfare (range 0-89). This measure is comprised of two subscales with 8-items each: Healthy behaviours (e.g., “My cat has been bright and alert”), and Clinical signs (e.g., “my cat has been ill or vomited”). Cronbach’s alphas: Total (α = .87), Healthy behaviours (α = .69), Clinical signs (α = .79).

#### Pet behavioural problems

For dog behaviour problems, The Mini C-BARQ (Canine Behavioural Assessment and Research Questionnaire) [83] was included. The measure is comprised of 42-items that examine owner perceptions of five key domains: Excitability, Aggression, Fear and anxiety, Separation-related issues, Attachment and attention seeking issues, Training and obedience difficulties, and Miscellaneous problems. Each item for each domain is scored on a severity scale of 1-4 (0 = No signs, to 4 = Severe signs), and frequency (i.e., 0 = Never, to 4 =Always). Positively worded items are reverse coded, and then total scores are calculated for each domain and across domains. Cronbach’s alphas: Excitability (α = .61), Aggression (α = .85), Fear and anxiety (α = .84), Separation-related behaviour (α = .73), Attachment and attention-seeking issues (α = .74), Training and obedience issues (α = .60), Miscellaneous problems (α = .75), Total (α = .89).

For cat behavioural problems, a measure of owner perceived cat behavioural problems was adapted from Grigg & Kogan [84] and is comprised of 9-items relating to perceived problematic behaviour (e.g., destructive behaviour, aggression, anxiety/fear, excessive vocalization, house soiling). Owners are asked to report Yes/No for whether their cat shows the specific behaviour, and total frequency scores are calculated (α = .50). As part of this measure, owners are also asked to rate the degree to which the behaviour (if relevant) ‘bothers them’, on 4-point scale from “Not bothered at all” to “Bothered a great deal”. A total score for how much the owner feels bothered by the problems is also calculated (α = .76).

#### Anxiety symptom severity

The Generalised Anxiety Disorder Questionnaire (GAD- 7) [85] is comprised of 7-items (e.g., “Not being able to stop or control worrying”, “Feeling afraid as if something awful might happen”). Participants are asked how often over the last two weeks they have been bothered by the symptoms. Each item is rated from 0 (Not at all) to 3 (Nearly every day). Total scores are calculated, providing a 0-21 severity score (α = .85). Scores of 5, 10, and 15 are taken as the cut-off points for mild, moderate, and severe anxiety, respectively. A score of 10 or greater represents a cut-off point for Generalized Anxiety Disorder.

#### Depression symptom severity

The depression module (PHQ-9) from the full PHQ (The Patient Health Questionnaire) [86] is comprised of 9-items (e.g., “Little interest or pleasure in doing things”, “Feeling down, depressed, or hopeless”). Participants are asked how often over the last two weeks they have been bothered by the symptoms. Each item is rated from 0 (Not at all) to 3 (Nearly every day). Total scores are calculated, providing a 0-27 severity score (α =.85). Cut-off points include scores of 0–4 for no depressive symptoms, 5–9 for mild depressive symptoms, 10–14 for moderate depressive symptoms, 15–19 for moderately-severe depressive symptoms, and 20–27 for severe depressive symptoms.

## Results

### Research question 1: Are there differences between dog and cat owners on measures of pet attachment and mental health?

First, we examined mental health symptom severity within our population. The majority of our participants met clinical cut offs for depression (74.5% of the sample) and anxiety (68.7% of the sample). Then, we examined whether there were differences between dog and cat owners on measures of mental health and pet attachment through independent t-tests. Although dog owners scored higher on anxiety and depression than cat owners, there were no significant differences found (both p>.05). Cat owners were more likely to display insecure attachments to their pets, but a significant difference was only found for anxious attachment (t(492)=-3.13, p=.002), and not avoidant attachment (p>.05). Descriptive statistics can be found in Table 2.

**Table 2.** Participant mental health symptom severity and pet attachment scores based on dog or cat ownership.

| **Depression** | **N (%) All** | **N (%) Dogs** | **N (%) Cats** |
| --- | --- | --- | --- |
| None | 28 (4.7) | 8 (2.3) | 20 (7.7) |
| Mild | 125 (20.8) | 80 (23.5) | 45 (17.4) |
| Moderate | 186 (31) | 101 (29.6) | 85 (32.8) |
| Moderate - severe | 152 (25.3) | 87 (25.5) | 65 (25.1) |
| Severe | 109 (18.2) | 65 (19.1) | 44 (17) |
| Mean (SD) | 13.65 (5.9) | 13.76 (5.7) | 13.51 (6.1) |
| **Anxiety** | **N (%) All** | **N (%) Dogs** | **N (%) Cats** |
| None | 32 (5.3) | 18 (5.3) | 14 (5.4) |
| Mild | 156 (26) | 90 (26.4) | 66 (25.5) |
| Moderate | 240 (40) | 127 (37.2) | 113 (43.6) |
| Severe | 172 (28.7) | 106 (31.1) | 66 (25.5) |
| Mean (SD) | 11.92 (4.7) | 12.07 (4.8) | 11.72 (4.5) |
| **Anxious pet attachment** | **Mean (SD), range All** | **Mean (SD), range Dogs** | **Mean (SD), range Cats** |
|  | 2.47 (.96), 1-5.88 | 2.36 (.86), 1-5 | 2.61 (1.05), 1-5.88 |
| **Avoidant pet attachment** | **Mean (SD), range All** | **Mean (SD), range Dogs** | **Mean (SD), range Cats** |
|  | 2.07 (.87), 1-6.25 | 2.04 (.85), 1-6.25 | 2.11 (.91), 1-5.38 |

*Notes:* ‘Moderate’ = cut off for both major depression and generalized anxiety disorder.

### Research question 2: Does insecure pet attachment relate to owner mental health symptom severity?

Next, we examined whether pet attachment predicted owner mental health severity through linear regressions (Table 3). For dog owners, anxious attachment significantly predicted higher severity scores for anxiety and depression. Therefore, dog owners who were anxiously attached were more likely to report higher anxiety and depression scores. For cat owners, avoidant attachment significantly predicted lower severity scores for anxiety and depression. Therefore, cat owners who displayed avoidant attachment were more likely to display lower anxiety and depression.

**Table 3.**
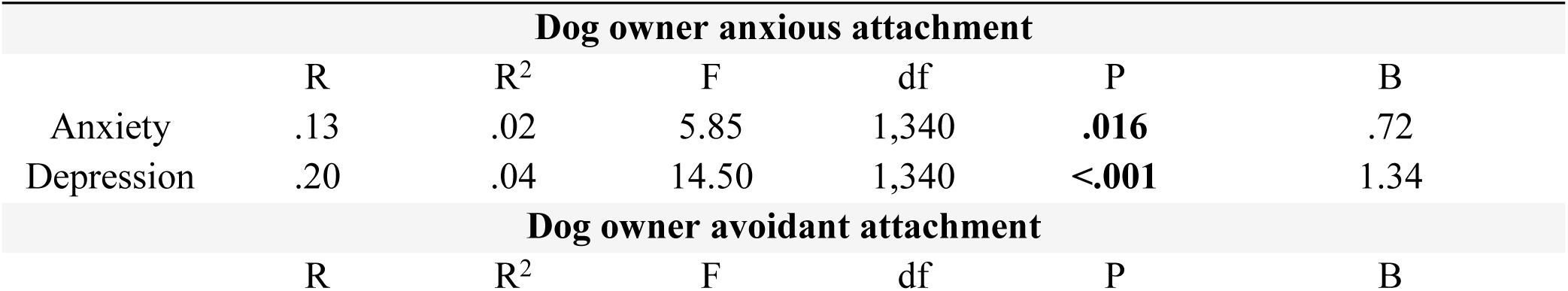
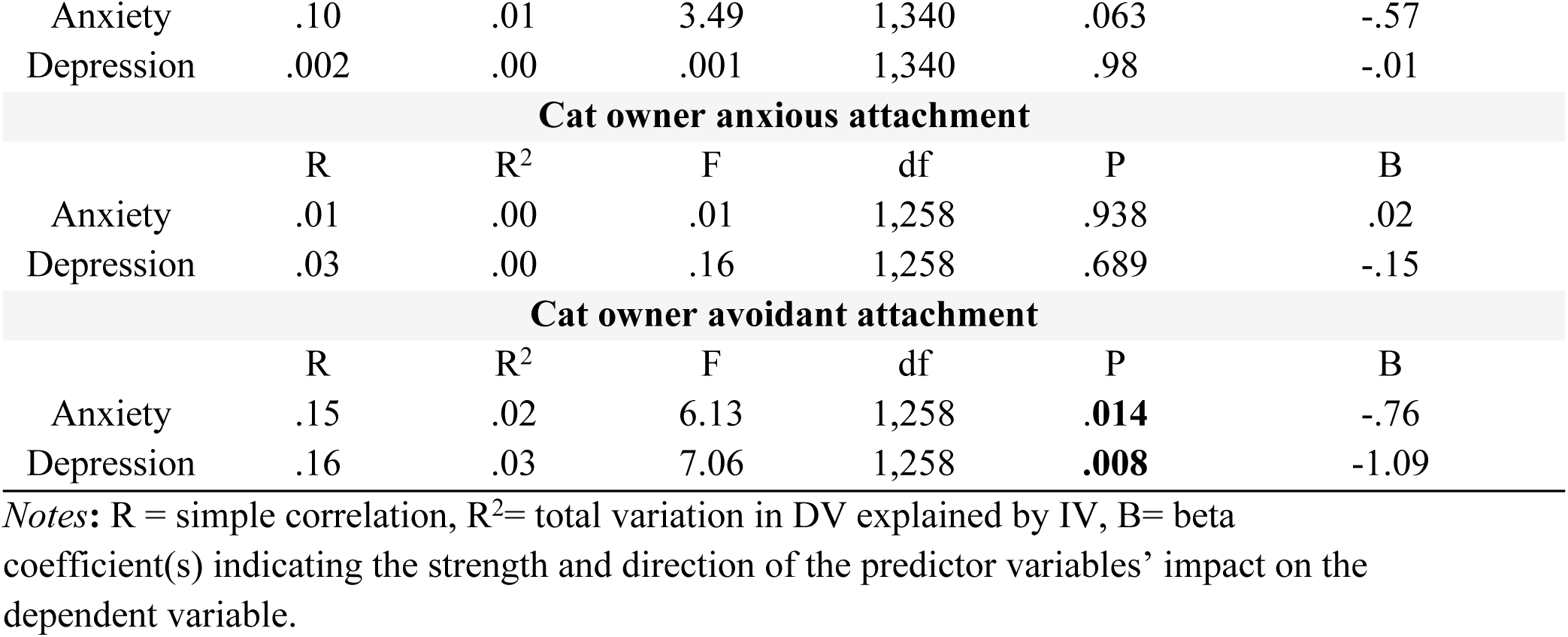
Linear regressions for insecure pet attachment predicting owner mental health severity.

| **Dog owner anxious attachment** |  |  |  |  |  |  |
| --- | --- | --- | --- | --- | --- | --- |
|  | R | R<sup>2</sup> | F | df | P | B |
| Anxiety | .13 | .02 | 5.85 | 1,340 | **.016** | .72 |
| Depression | .20 | .04 | 14.50 | 1,340 | **<.001** | 1.34 |
| **Dog owner avoidant attachment** |  |  |  |  |  |  |
|  | R | R<sup>2</sup> | F | df | P | B |
| Anxiety | .10 | .01 | 3.49 | 1,340 | .063 | -.57 |
| Depression | .002 | .00 | .001 | 1,340 | .98 | -.01 |
| **Cat owner anxious attachment** |  |  |  |  |  |  |
|  | R | R<sup>2</sup> | F | df | P | B |
| Anxiety | .01 | .00 | .01 | 1,258 | .938 | .02 |
| Depression | .03 | .00 | .16 | 1,258 | .689 | -.15 |
| **Cat owner avoidant attachment** |  |  |  |  |  |  |
|  | R | R<sup>2</sup> | F | df | P | B |
| Anxiety | .15 | .02 | 6.13 | 1,258 | **.014** | -.76 |
| Depression | .16 | .03 | 7.06 | 1,258 | **.008** | -1.09 |

Notes: R = simple correlation, R<sup>2</sup>= total variation in DV explained by IV, B= beta coefficient(s) indicating the strength and direction of the predictor variables' impact on the dependent variable.

### Research question 3: Does perceived pet welfare explain the relationship between insecure pet attachment and owner mental health symptom severity?

To test assumptions for mediation analysis, we examined relationships between 1) pet attachment and mental health, 2) pet attachment and perceived pet welfare, and 3) perceived pet welfare and owner mental health. First, we examined the relationship between pet attachment and owner mental health (see Tables 4, 5). For dog owners, attachment anxiety significantly positively correlated with both anxiety and depression scores. Attachment avoidance significantly negatively correlated with anxiety scores. For cat owners, attachment avoidance significantly negatively correlated with both anxiety and depression.

**Table 4.**
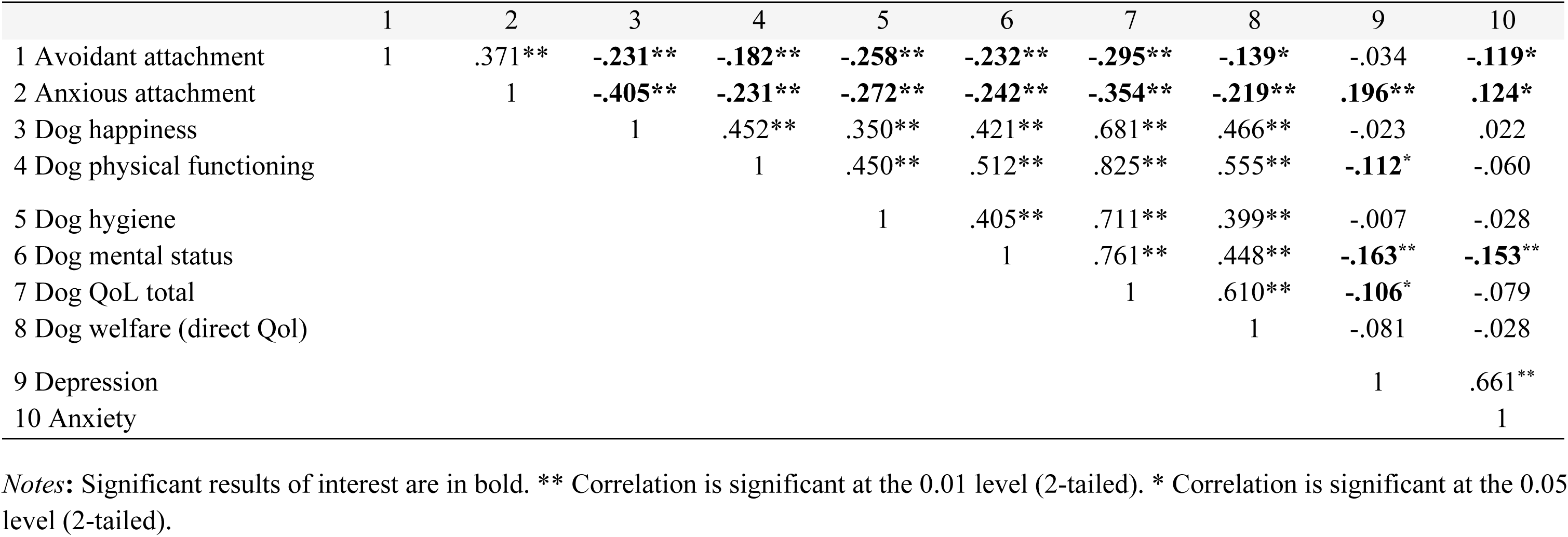
Spearman two-tailed correlations between pet attachment and perceived dog welfare, and between perceived dog welfare and owner mental health.

**Table 5.** Spearman two-tailed correlations between pet attachment and perceived cat welfare, and between perceived cat welfare and owner mental health.

|  | 1 | 2 | 3 | 4 | 5 | 6 | 7 | 8 |
| --- | --- | --- | --- | --- | --- | --- | --- | --- |
| 1 Avoidant attachment | 1 | .315** | **-.365\*\*** | **-.163\*\*** | **-.294\*\*** | **-.159\*** | **-.177\*\*** | **-.190\*\*** |
| 2 Anxious attachment |  | 1 | **-.385\*\*** | **-.275\*\*** | **-.375\*\*** | **-.321\*\*** | -.034 | -.008 |
| 3 Cat healthy behaviours |  |  | 1 | **.381\*\*** | **.791\*\*** | **.387\*\*** | .093 | .096 |
| 4 Cat clinical signs |  |  |  | 1 | **.793\*\*** | **.406\*\*** | -.018 | .007 |
| 5 Cat QoL total |  |  |  |  | 1 | **.478\*\*** | .024 | .039 |
| 6 Cat welfare (direct QoL assessment) |  |  |  |  |  | 1 | .045 | **.132\*** |
| 7 Depression |  |  |  |  |  |  | 1 | .715** |
| 8 Anxiety |  |  |  |  |  |  |  | 1 |

*Notes:* significant results of interest are in bold. \*\* Correlation is significant at the 0.01 level (2-tailed). \* Correlation is significant at the 0.05 level (2-tailed).

For dog owners, we tested correlations between attachment, perceived pet quality of life (direct assessment scale), individual subscales and total scores on the CHQLS-15, and owner mental health (see Table 4). Anxious and avoidant attachment were both significantly negatively associated with all dog welfare outcomes. Therefore, dog owners who were more insecurely attached, were more likely to rate their dog’s quality of life and welfare lower. Scores on the subscale ‘physical functioning’ were significantly negatively associated with owner depression scores, i.e. better dog physical functioning was associated with lower depression. Scores on the subscale ‘mental status’ were significantly negatively associated with both anxiety and depression, i.e. better dog mental status (e.g., happier) were associated with lower anxiety and depression. Total scores on the CHQLS-15 were significantly negatively associated with depression, i.e. higher pet welfare was associated with lower depression.

For cat owners, we tested correlations between attachment, perceived pet quality of life (direct assessment scale), subscales and total scores on the FHQLS, and owner mental health (see Table 5). There was a significant negative correlation between both avoidant and anxious attachment, and the two subscales of the FHQLS, total FHQLS, and cat welfare (direct QoL). Therefore, cat owners who were insecurely attached were more likely to rate their cats’ welfare as lower. There was a significant positive correlation between cat welfare (direct QoL assessment) and anxiety, i.e. higher perceived cat welfare was associated with higher anxiety. No other correlations were significant.

Next, variables which met required assumptions (i.e. interrelationships existed between them) were further analysed with mediation analysis. First, we examined the mediational effect of a dog’s physical functioning (M) on the relationship between anxious attachment (X) and owner depression (Y), and the relationship between avoidant attachment (X) and owner anxiety (Y) (Table 6). No significant mediations were found; the indirect effects were not significant.

**Table 6.** Parallel mediation analysis examining a) indirect effects of anxious owner-dog attachment (X) on depression symptom severity (Y), via dogs physical functioning (M), and b) indirect effects of avoidant owner-dog attachment (X) on anxiety symptom severity (Y), via dogs physical functioning (M).

|  | Indirect effects of anxious owner-dog attachment (X)<br>on depression symptom severity (Y), via dogs<br>physical functioning (M). |  |  | Indirect effects of avoidant owner-dog attachment<br>(X) on anxiety symptom severity (Y), via dogs<br>physical functioning (M). |  |  |
| --- | --- | --- | --- | --- | --- | --- |
| | $\beta$ | SE | 95% CI | $\beta$ | SE | 95% CI |
| Completely standardised<br>indirect effect beta values of<br>X on Y ($ab_{cs}$) (total) | .016 | .012 | -.005, .040 | .018 | .012 | -.001, .044 |
| Direct effect of X on M ($a_1$) | -.149* | .039 | -.225, -.073 | -.142* | .040 | -.220, -.064 |
| Direct effect of M on Y ($b_1$) | -.703 | .494 | -1.674, .268 | -.708 | .420 | -1.530, .115 |
| Direct effect of X on Y ($c'$) | 1.236* | .359 | .529, 1.943 | -.670* | .310 | -1.280, -.060 |
| Indirect effect of X on Y via<br>M | .105 | .077 | -.036, .271 | .101 | .066 | -.005, .247 |

*Notes:* \* Significant pathway ( $p < 0.05$ ). Effect sizes: $abcs = 0.01$ (small effect), $abcs = 0.09$ (medium effect), and $abcs = 0.25$ (large effect). M= dogs physical functioning.

Next, we examined the mediational effect of a dog’s mental status (M) on the relationship between anxious attachment (X) and owner anxiety and depression (Y), and on the relationship between avoidant attachment (X) and owner anxiety (Y) (Table 7, Figure 1). Owner-dog anxious attachment had a significant indirect effect on owner anxiety symptom severity through dog’s mental status (abcs=.026, large effect); this was a complete mediation as the direct effect of X on Y was no longer significant when accounting for M. Owner-dog anxious attachment had a significant indirect effect on owner depression symptom severity through dog’s mental status (abcs=.028, large effect); this was a partial mediation as the direct effect of X on Y remained significant (p=.001) when accounting for M. Owner-dog avoidant attachment had a significant indirect effect on owner anxiety symptom severity through dog’s mental status (abcs=.055, large effect); this was a partial mediation as the direct effect of X on Y remained significant (p=.001) when accounting for M.

**Figure 1.**
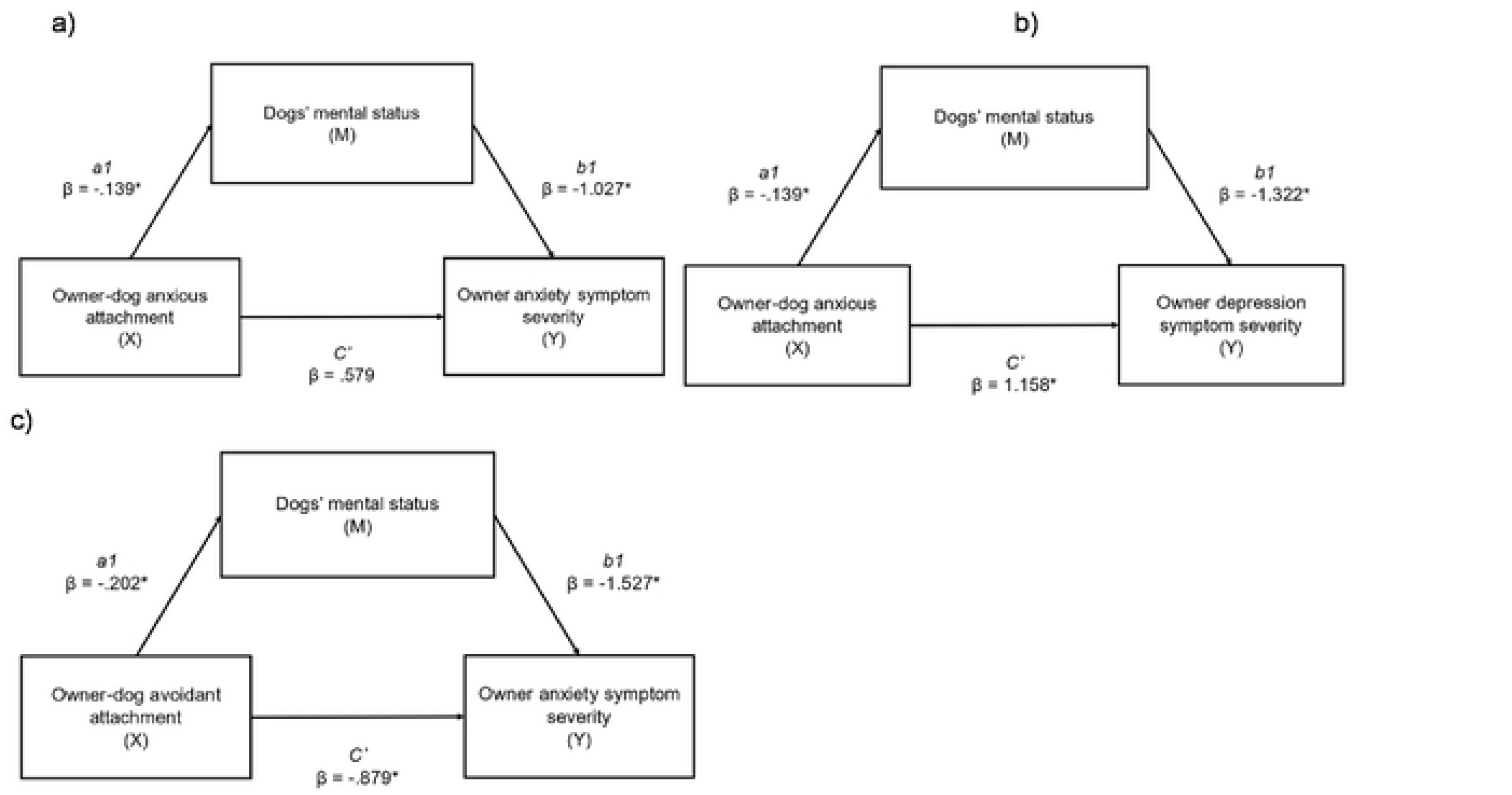
a) indirect effects of anxious owner-dog attachment (X) on anxiety symptom severity (Y), viadogs mental status (M). (abcs=.026, largeeffect); b) indirect effects of anxious owner-dog attachment (X) on depression symptom severity (Y), via dogs mental status (M). (abcs=.028, large effect); c) indirect effects of avoidant owner-dog attachment (X) on anxiety symptom severity (Y), via dogs mental status (M). (abcs=.055, large effect). *=significant pathway.

**Table 7.**
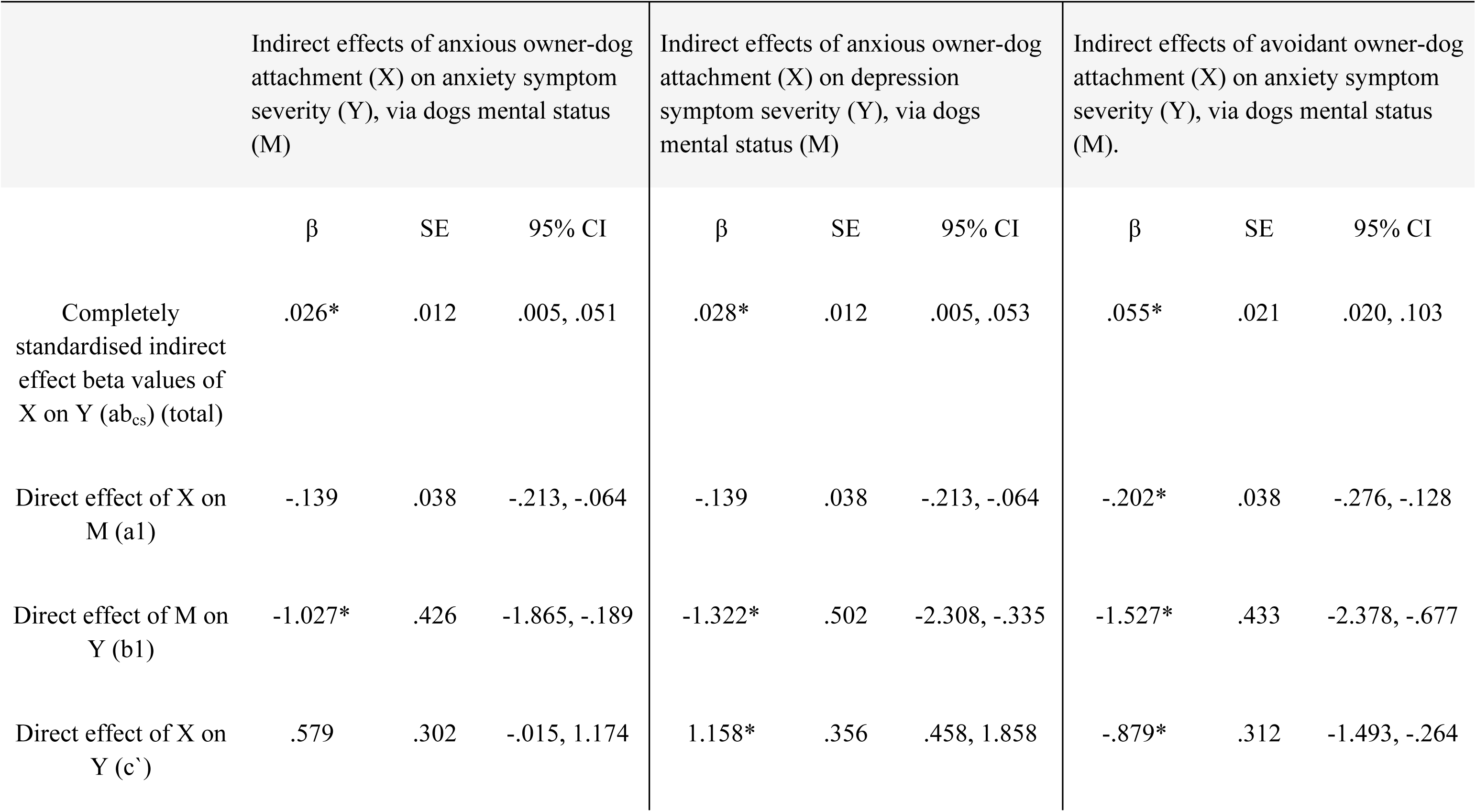
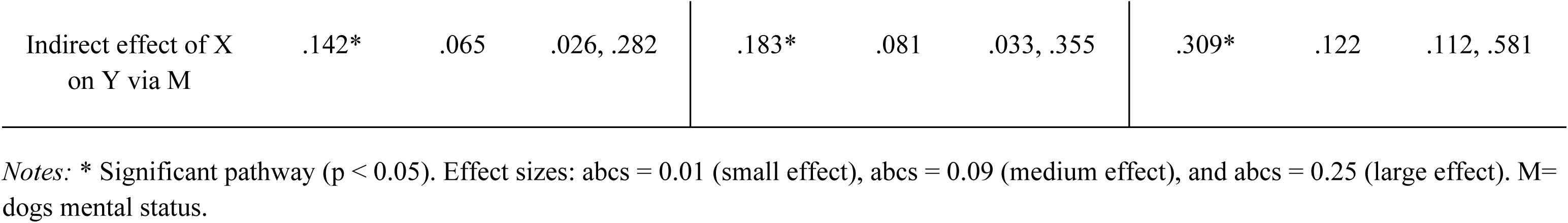
Parallel mediation analysis examining a) indirect effects of anxious owner-dog attachment (X) on anxiety symptom severity (Y), via dogs mental status (M), and b) parallel mediation analysis examining a) indirect effects of anxious owner-dog attachment (X) on depression symptom severity (Y), via dogs mental status (M), and c) indirect effects of avoidant owner-dog attachment (X) on anxiety symptom severity (Y), via dogs mental status (M).

|  | Indirect effects of anxious owner-dog attachment (X) on anxiety symptom severity (Y), via dogs mental status (M) |  |  | Indirect effects of anxious owner-dog attachment (X) on depression symptom severity (Y), via dogs mental status (M) |  |  | Indirect effects of avoidant owner-dog attachment (X) on anxiety symptom severity (Y), via dogs mental status (M). |  |  |
| --- | --- | --- | --- | --- | --- | --- | --- | --- | --- |
| | $\beta$ | SE | 95% CI | $\beta$ | SE | 95% CI | $\beta$ | SE | 95% CI |
| Completely standardised indirect effect beta values of X on Y ($ab_{cs}$) (total) | .026* | .012 | .005, .051 | .028* | .012 | .005, .053 | .055* | .021 | .020, .103 |
| Direct effect of X on M (a1) | -.139 | .038 | -.213, -.064 | -.139 | .038 | -.213, -.064 | -.202* | .038 | -.276, -.128 |
| Direct effect of M on Y (b1) | -1.027* | .426 | -1.865, -.189 | -1.322* | .502 | -2.308, -.335 | -1.527* | .433 | -2.378, -.677 |
| Direct effect of X on Y (c') | .579 | .302 | -.015, 1.174 | 1.158* | .356 | .458, 1.858 | -.879* | .312 | -1.493, -.264 |
| Indirect effect of X<br>on Y via M | .142* | .065 | .026, .282 | .183* | .081 | .033, .355 | .309* | .122 | .112, .581 |

Notes: \* Significant pathway ( $p < 0.05$ ). Effect sizes: $abcs = 0.01$ (small effect), $abcs = 0.09$ (medium effect), and $abcs = 0.25$ (large effect). M=
dogs mental status.

Next, we examined the mediational effect of a dog’s welfare (total CHQLS) (M) on the relationship between anxious attachment (X) and owner depression (Y), and the mediational effect of a cat’s welfare (direct QoL assessment) (M) on the relationship between avoidant attachment (X) and owner anxiety (Y) (Table 8). No significant mediations were found; the indirect effects were not significant.

**Table 8.** Mediation analysis examining a) indirect effects of anxious owner-dog attachment (X) on depression symptom severity (Y), via dogs total quality of life scores (CHQLS) (M), and b) indirect effects of avoidant owner-cat attachment (X) on anxiety symptom severity (Y), via cats quality of life scores (direct assessment) (M).

|  | Indirect effects of anxious owner-dog attachment (X)<br>on depression symptom severity (Y), via dogs total<br>quality of life scores (CHQLS) (M). |  |  | Indirect effects of avoidant owner-cat attachment (X)<br>on anxiety symptom severity (Y), via cats quality of<br>life scores (direct assessment) (M). |  |  |
| --- | --- | --- | --- | --- | --- | --- |
| | $\beta$ | SE | 95% CI | $\beta$ | SE | 95% CI |
| Completely standardised<br>indirect effect beta values of<br>X on Y ($ab_{cs}$) (total) | .018 | .018 | -.019, .053 | .018 | .022 | -.023, .063 |
| Direct effect of X on M (a1) | -.160* | .026 | -.212, -.108 | -3.080* | .562 | -4.186, -1.974 |
| Direct effect of M on Y (b1) | -.763 | .728 | -2.195, .699 | -.029 | .034 | -.096, .039 |
| Direct effect of X on Y (c') | 1.219* | .371 | .489, 1.948 | -.853* | .326 | -1.495, -.211 |
| Indirect effect of X on Y via<br>M | .122 | .121 | -.127, .356 | .089 | .107 | -.117, .313 |

*Notes:* \* Significant pathway ( $p < 0.05$ ). Effect sizes: $abcs = 0.01$ (small effect), $abcs = 0.09$ (medium effect), and $abcs = 0.25$ (large effect). M= pet quality of life.

### Research question 4: Do perceived pet behavioural problems explain the relationship between insecure attachment and owner mental health symptom severity?

To test assumptions for mediation analysis, we first examined correlations between owner-pet attachment, perceived pet behavioural problems, and owner mental health. For dog owners, we tested correlations between owner-dog attachment, perceived dog behavioural problems using both totals and subscales of the CBARQ, and owner mental health (Table 9). There was a significant negative relationship between owner-dog avoidant attachment and dog excitability, and attachment related issues, and a significant positive relationship between owner-dog avoidant attachment and dog aggression and training. Therefore, avoidant attached dog owners scored their dogs lower on excitability and attachment related issues and scored their dog higher on aggression and training difficulty. There was a significant negative relationship between owner-dog anxious attachment and dog excitability, and a significant positive relationship between owner-dog anxious attachment and all other subscales (except for attachment related issues) and total issues. Therefore, dog owners who scored high on anxious attachment were more likely to report more dog behavioural problems.

**Table 9.** Relationships between owner-pet attachment, perceived pet behavioural problems, and owner mental health.

|  |  | Avoidant<br>attachment | Anxious<br>attachment | Owner<br>depression | Owner<br>anxiety |
| --- | --- | --- | --- | --- | --- |
| Dog owners | Excitability | -.147** | -.107* | -0.014 | 0.033 |
|  | Aggression | .189** | .182** | 0.078 | 0.043 |
|  | Fear & Anxiety | 0.012 | .157** | .165** | .146** |
|  | Separation issues | 0.037 | .190** | 0.085 | 0.063 |
|  | Attachment related issues | -.204** | -0.099 | 0.088 | 0.093 |
|  | Training difficulty | .160** | .238** | .146** | 0.041 |
|  | Misc | 0.106 | .133* | 0.063 | 0.036 |
|  | Total issues | 0.032 | .148* | 0.067 | 0.075 |
| Cat owners | Cat behavioural problems (total) | 0.005 | .123* | -0.012 | -0.01 |
|  | Ratings of being 'bothered' by problems | 0.308 | .409* | 0.158 | 0.013 |

*Notes:* significant results of interest are in bold. \*\* Correlation is significant at the 0.01 level (2-tailed). \* Correlation is significant at the 0.05 level (2-tailed).

There was a significant positive relationship between scores for dogs’ fear and anxiety and owner symptoms of anxiety and depression. There was a significant positive relationship between dog training difficulty and owner depression. Therefore, owners who report more dog behavioural problems (in this case, only fear and anxiety) scored higher on anxiety and depression symptom severity.

For cat owners, we examined both the total behavioural problems measure and owners’ ratings for ‘how bothered’ they feel by such problems (Table 9). There was a significant positive relationship between anxious owner-cat attachment and perceived cat behavioural problems and a significant positive relationship between anxious owner-cat attachment and ratings of being ‘bothered’ by such problems; thus, anxious cat owners were more likely to report more cat behavioural problems and were more likely to report being bothered by these problems. These findings were not replicated for avoidant attachment.

Variables which met the required assumptions were further tested with mediation analysis. First, we examined the mediational effect of a dog’s fear and anxiety (M) on the relationship between anxious attachment (X) and owner anxiety (Y), and on the relationship between anxious attachment (X) and owner depression (Y) (Table 10, Figure 2). Owner-dog anxious attachment had a significant indirect effect on owner anxiety symptom severity through dog’s fear and anxiety (abcs=.020, medium effect); this was a partial mediation as the direct effect of X on Y remained significant (p=.001) when accounting for M. Owner-dog anxious attachment had a significant indirect effect on owner depression symptom severity through dog’s fear and anxiety (abcs=.021, medium effect); this was a partial mediation as the direct effect of X on Y remained significant (p=.001) when accounting for M.

**Figure 2.**
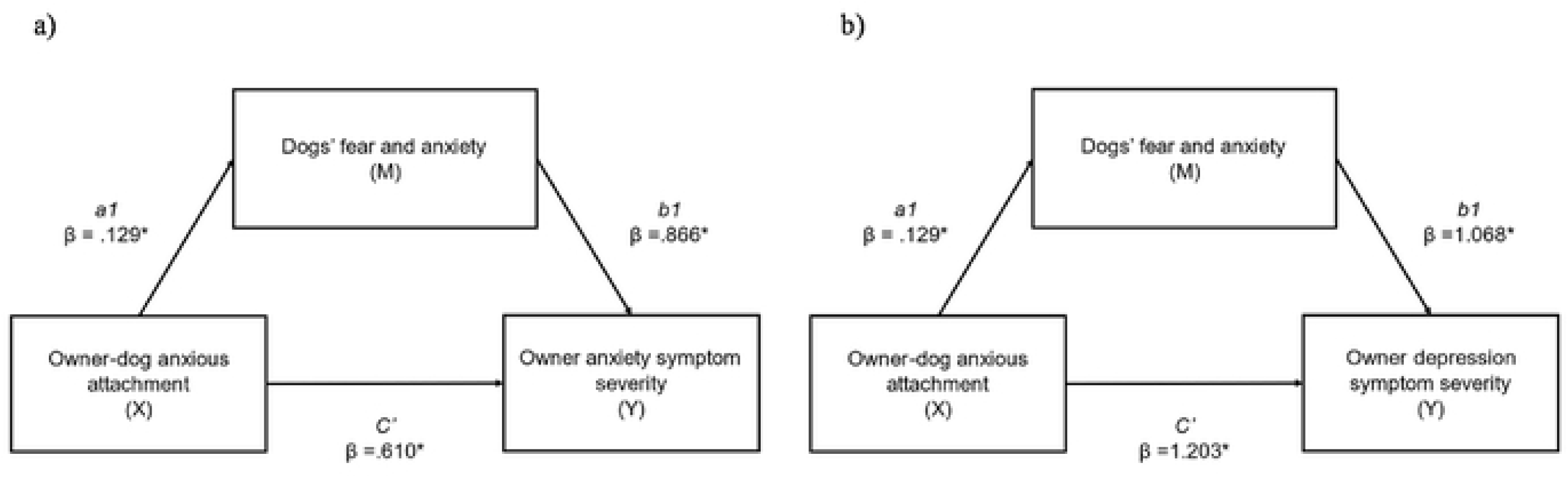
a) indirect effects of anxious owner-dog attachment (X) on anxiety symptom severity (Y), via dogs fear and anxiety (M). (abcs=.020, medium effect); b) indirect effects of anxious owner.dog attachment (X) on depression symptom severity (Y). via dogs fear and anxiety (M). (abcs=.021, medium effect). *=significant pathway.

**Table 10.** Mediation analysis examining a) indirect effects of anxious owner-dog attachment (X) on anxiety symptom severity (Y), via dog fear and anxiety (M), and b) indirect effects of anxious owner-dog attachment (X) on depression symptom severity (Y), via dog fear and anxiety (M), and c) indirect effects of anxious owner-dog attachment (X) on depression symptom severity (Y), via dog training difficulty (M).

|  | Anxious owner-dog attachment (X)<br>on anxiety symptom severity (Y),<br>via dog fear and anxiety (M). |  |  | Anxious owner-dog attachment (X)<br>on depression symptom severity (Y),<br>via dog fear and anxiety (M). |  |  | Anxious owner-dog attachment (X)<br>on depression symptom severity (Y),<br>via dog training difficulty (M). |  |  |
| --- | --- | --- | --- | --- | --- | --- | --- | --- | --- |
| | $\beta$ | SE | 95% CI | $\beta$ | SE | 95% CI | $\beta$ | SE | 95% CI |
| Completely<br>standardised indirect<br>effect beta values of<br>X on Y ($ab_{cs}$) (total) | .020* | .068 | .012, .273 | .021* | .012 | .002, .050 | .026 | .015 | -.001, .057 |
| Direct effect of X on<br>M (a1) | .129* | .049 | .033, .224 | .129* | .049 | .033, .224 | .196* | .043 | .112, .281 |
| Direct effect of M on<br>Y (b1) | .866* | .331 | .215, 1.517 | 1.068* | .390 | .301, 1.835 | .872* | .442 | .003, 1.742 |
| Direct effect of X on<br>Y (c') | .610* | .299 | .022, 1.199 | 1.203* | .352 | .510, 1.897 | 1.170* | .361 | .459, 1.880 |
| Indirect effect of X<br>on Y via M | .111* | .068 | .012, .273 | .137* | .084 | .016, .334 | .171 | .097 | -.006, .384 |

Notes: \* Significant pathway ( $p < 0.05$ ). Effect sizes: $abcs = 0.01$ (small effect), $abcs = 0.09$ (medium effect), and $abcs = 0.25$ (large effect). M= dogs fear and anxiety.

We then examined the mediational effect of dog training difficulty (M) on the relationship between owner-dog anxious attachment (X) and depression (Y) (Table 10). No significant mediation was found; the indirect effect was not significant. A significant positive direct effect was however found for dogs’ training difficulty on owner depression; thus, those who reported more difficulties with training their dog, also reported higher depression.

### Research question 5: Does owner-pet compatibility explain the relationship between insecure attachment and owner mental health symptom severity?

To test assumptions for mediation analysis, we first examined correlations between owner-pet attachment, perceived owner-pet compatibility, and owner mental health (Table 11). There were significant negative relationships between owner-dog avoidant attachment and all compatibility subscales (except for physical) and total compatibility scores; thus, those who scored high on avoidant attachments felt less compatible with their dogs. There were significant negative relationships between owner-dog anxious attachment and all compatibility subscales and total compatibility scores; thus, those with anxious attachments felt less compatible with their dogs. There was a significant positive relationship between the affection subscale and owner anxiety; thus, those who reported higher compatibility in the affection domain, scored higher on anxiety.

**Table 11.** Relationships between owner-pet attachment and owner-pet compatibility (subscales and total score), and between owner-pet compatibility and owner mental health.

| Dog owners | Physical | Social | Affection | Closeness | Other | Total |
| --- | --- | --- | --- | --- | --- | --- |
| Owner-dog attachment avoidance | -.098 | **-.129\*** | **-.325\*\*** | **-.375\*\*** | **-.207\*\*** | **-.382\*\*** |
| Owner-dog attachment anxiety | **-.158\*\*** | **-.086** | **-.269\*\*** | **-.285\*\*** | **-.107\*** | **-.308\*\*** |
| Owner depression | -.045 | -.022 | .065 | -.023 | .026 | .001 |
| Owner anxiety | -.029 | -.081 | **.149\*\*** | .074 | .075 | .065 |
| Cat owners | Physical | Social | Affection | Closeness | Other | Total |
| Owner-cat attachment avoidance | .052 | -.052 | **-.277\*\*** | **-.252\*\*** | -.086 | **-.246\*\*** |
| Owner-cat attachment anxiety | -.044 | **-.225\*\*** | **-.246\*\*** | **-.273\*\*** | -.035 | **-.306\*\*** |
| Owner depression | .095 | .021 | .0001 | .113 | .957 | .197 |
| Owner anxiety | .028 | .014 | -.008 | .113 | .026 | .062 |

*Notes:* significant results of interest are in bold. \*\* Correlation is significant at the 0.01 level (2-tailed). \* Correlation is significant at the 0.05 level (2-tailed).

These analyses were then replicated for cat owners (Table 11). There were significant negative relationships between owner-cat avoidant attachment, subscales for affection and closeness, and total compatibility scores; thus, those with avoidant attachments felt less compatible with their cats. There were significant negative relationships between owner-cat anxious attachment, and all subscales (except physical and other), and total compatibility scores; thus, those with anxious attachments felt less compatible with their cats. There were no significant relationships between owner-cat compatibility and owner mental health.

Next, variables which met required assumptions (i.e. interrelationships existed between them) were further analysed with mediation analysis. First, we examined the mediational effect of affection compatibility (M) on the relationship between owner-dog anxious attachment (X) and owner anxiety (Y), and on the relationship between owner-dog avoidant attachment (X) and owner anxiety (Y) (Table 12, Figure 3). Owner-dog anxious attachment had a significant indirect effect on owner anxiety symptom severity through affection compatibility (abcs=-.053, large effect); this was a partial mediation as the direct effect of X on Y remained significant when accounting for M. Owner-dog avoidant attachment had a significant indirect effect on owner anxiety symptom severity through affection compatibility (abcs= -.042, large effect); this was a complete mediation as the direct effect of X on Y was no longer significant when accounting for M.

**Figure 3.**
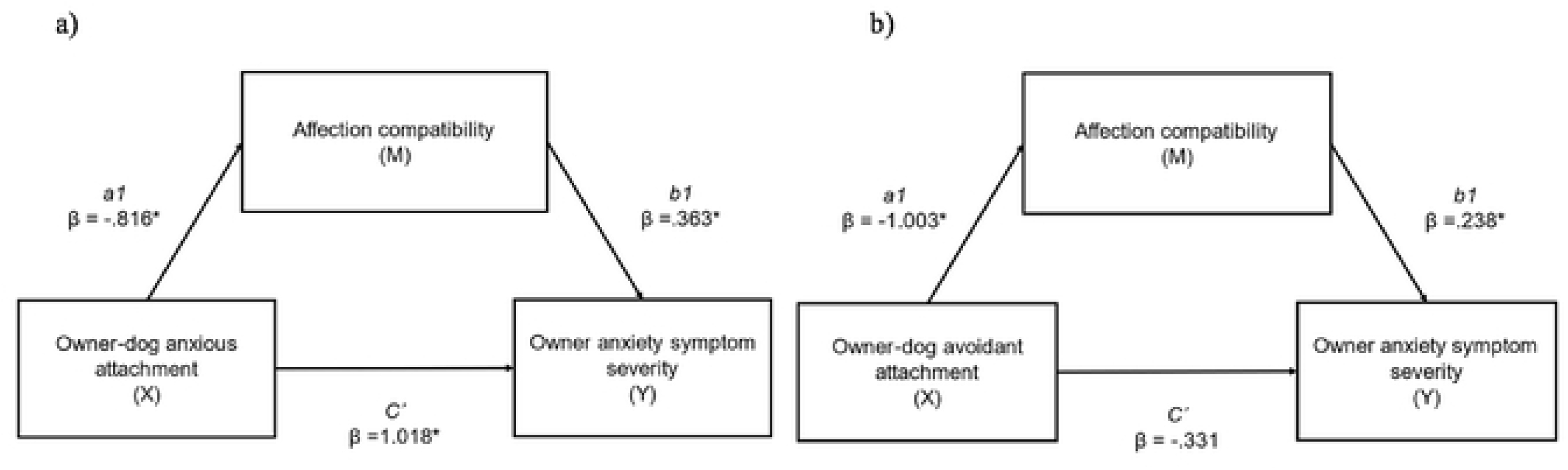
a) indirect cfTocts of anxious owner-dog attachment (X) on anxiety symptom severity (Y), via alTection compatibility (M). (abcs=­ .053, large effect); b) indirect effects of avoidant owner-dog attachment (X) on anxiety symptom severity (Y), via affection compatibility (M). (abcs=-.042, large effect). *=significant pathway.

**Table 12.** Mediation analysis examining a) indirect effects of anxious owner-dog attachment (X) on anxiety symptom severity (Y), via affection compatibility (M), and b) indirect effects of avoidant owner-dog attachment (X) on anxiety symptom severity (Y), via affection compatibility (M).

|  | Indirect effects of anxious owner-dog attachment (X)<br>on anxiety symptom severity (Y), via affection<br>compatibility (M). |  |  | Indirect effects of avoidant owner-dog attachment<br>(X) on anxiety symptom severity (Y), via affection<br>compatibility (M). |  |  |
| --- | --- | --- | --- | --- | --- | --- |
| | $\beta$ | SE | 95% CI | $\beta$ | SE | 95% CI |
| Completely standardised<br>indirect effect beta values of X<br>on Y ($ab_{cs}$) (total) | -.053* | .018 | -.912, -.021 | -.042* | .020 | -.083, -.005 |
| Direct effect of X on M (a1) | -.816* | .159 | -1.129, -.504 | -1.003* | .159 | -1.316, -.691 |
| Direct effect of M on Y (b1) | .363* | .100 | .165, .560 | .238* | .104 | .034, .442 |
| Direct effect of X on Y (c') | 1.018* | .305 | .419, 1.617 | -.331 | .321 | -.961, .299 |
| Indirect effect of X on Y via M | -.296* | .100 | -.502, -.118 | -.239* | .112 | -.465, -.026 |

*Notes:* \* Significant pathway ( $p < 0.05$ ). Effect sizes: $abcs = 0.01$ (small effect), $abcs = 0.09$ (medium effect), and $abcs = 0.25$ (large effect). M= affection compatibility.

## Discussion

This study aimed to examine whether the relationship between owner-pet attachment and owner mental health, can be explained by owner perceived pet compatibility, pet welfare, and pet behavioural problems. These aspects, often overlooked in previous research, are believed to play crucial roles in shaping owner-pet relationships and consequently influencing owner mental wellbeing. This study focused on emerging young adults within the United Kingdom who self-identified as having mental health difficulties. While our study sample was drawn from the general population, it’s noteworthy that a significant proportion of the young adults recruited met clinical thresholds for Generalized Anxiety Disorder and/or Major Depressive Disorder, and many exhibited co-morbidity and additional diagnoses. Therefore, our study focused on an underrepresented population within this field of study, and it important to consider the population when interpreting the findings of this study.

Firstly, our findings revealed no significant differences in mental health symptom severity between dog and cat owners, which could be explained by the high rates of symptoms in both types of owners within our sample. There were differences, however, between dog and cat owners on attachment scores. Dog owners scored higher on pet attachment security compared to cat owners, who exhibited higher attachment anxiety scores. While cat owners also scored higher on avoidant attachment scores, this disparity did not reach statistical significance. These results align with previous research, which has demonstrated that cat owners tend to display more insecure attachments and emotional distance in their pet relationships compared to dog owners who tend to display the reverse [41, 87, 88]. These findings could possibly be explained by species-typical differences in social behaviour. Individuals with avoidant attachment tendencies may seek independence and autonomy in relationships, which could align more closely with the characteristics of a less physically and emotionally demanding pet, one which displays more avoidant attachment-related characteristics, and one in which there are lower expectations from the relationship, such as a cat [67, 89, 90]. However, this hypothesis fails to fully account for why cat owners also exhibited higher levels of anxiety in their pet relationships. Anxious attachment typically entails an increased need for reassurance and emotional closeness from relationships, characteristics that might be more readily fulfilled by dogs, given their greater dependence and reliance on their owners [66]. Interestingly, Beck & Madresh [28] also found higher relationship anxiety in cat owners and proposed that cat ownership could be a response to loneliness. Further research is needed into attachment-related differences between dog and cat owners before any firm conclusions can be drawn. Qualitative investigations could explore individuals’ experiences and preferences regarding specific pet species and individual characteristics, considering their attachment orientations. Future research could also expand its scope to include an individual’s broader interpersonal relationships, investigating whether attachment orientations observed in human relationships extend to those with pets, or whether pet attachments have unique qualities that can buffer against a lack of secure human attachment, potentially mitigating psychological distress.

Previous hypotheses suggest that individuals facing mental health difficulties may be more inclined to acquire a pet and feel more closely bonded to their pets, actively seeking out pets for emotional support and comfort as a strategy for managing their wellbeing. This notion could potentially shed light on the inconsistencies observed in the field regarding the purported beneficial impacts of pets on mental health [29, 91]. However, the reverse has also been proposed whereby secure pet attachment can be a protective factor for mental health difficulties [41, 92]. We found partial support for both hypotheses; higher attachment avoidance scores predicted lower anxiety scores regardless of pet species, whereas higher attachment anxiety scores predicted higher depression scores for dog owners only. The findings for anxious pet attachment align with both human-human and human-pet attachment research, indicating that insecure attachment can potentially contribute to poorer mental health outcomes [41, 93], with larger associations found for attachment anxiety [94]. Conversely, the unexpected findings concerning avoidant pet attachment suggest that avoidance might serve as a protective factor against pet owner anxiety. It could be posited that avoidantly attached individuals display fears of rejection judgement in their pet relationships in a similar way observed within their close human relationships. In this context, pets can offer a unique form of unconditional and non-judgemental support and acceptance which may serve as a protective buffer against diminished wellbeing [17, 23]. Pets can also provide a secure and non-judgmental avenue for emotional expression and can facilitate emotion regulation for avoidantly attached individuals who may encounter challenges in expressing and regulating their emotions in interpersonal relationships [62, 95]. This could be particularly important for our emerging adult sample who display high mental health symptom severity. This life stage is associated with significant transitions and uncertainty, and societal, psychosocial, and biological factors which can increase psychological distress [5]. These theories need further testing to fully understand the relationship between attachment orientations and mental health outcomes in the context of pet ownership.

We also explored whether perceived pet welfare explained the relationship between insecure pet attachment and owner mental health symptom severity. Our findings indicated that insecurely attached (anxious and avoidant) dog and cat owners were more likely to rate their pet’s quality of life lower. An individual’s attachment system and caregiving system operate in tandem [39], suggesting that owner-pet attachment could influence how owners perceive and interact with their pets. Previous research supports this theory, demonstrating that stronger pet attachment relates to increased caregiving behaviour and positive owner-pet interactions across the lifespan, potentially improving welfare outcomes for pets [61, 62]. However, it has been proposed that anxious attachments may in fact foster greater care and attentiveness, whereas avoidant individuals might exhibit more neglectful behaviours [66, 67]. It is important to note that our findings may reflect owner perceptions of their pet’s quality of life rather than the actual welfare of their animals. Insecurely attached individuals often have internal working models that view interpersonal relationships more negatively, leading to heightened worries, concerns, sensitivity to rejection, and challenges in accurately interpreting emotional cues [63, 65]. These tendencies might manifest in certain emotional responses to pets, such as attributing negative emotions to them, or misinterpreting ambiguous pet behaviour as distress or discomfort, thereby potentially perceiving their pet’s welfare as lower, even without substantiated evidence. This theory finds support in qualitative research where individuals with heightened anxiety tend to report maladaptive stress, worries, and anxiety over their pet’s welfare. These individuals tend to express feelings of rejection when a pet fails to meet their expectations in a particular interaction, such as not reciprocating physical affection in a time of need [23, 25]. Moreover, insecurely attached individuals who perceive themselves as being unable to, or are not, currently meeting their pet’s welfare needs, may experience feelings of failure, intensifying feelings of insecurity, and exacerbating mental health symptom severity [23, 96].

Regarding perceived pet welfare and owner mental health, for dog owners, higher total welfare scores correlated with reduced depression, while elevated scores on positive physical functioning were linked to decreased anxiety. Additionally, higher scores on positive mental status were associated with lower levels of both anxiety and depression. These findings are in support of past research highlighting that poor pet welfare can be a risk factor for poor mental wellbeing in owners [24, 97]. These findings were not replicated in cat owners within our study. We also found no mediational effect of a dog or cat’s total quality of life scores on the relationship between attachment and owner mental health. However, a noteworthy finding emerged regarding a complete mediational effect for a dog’s mental status. Dog owners who scored higher on attachment insecurity (both anxious and avoidant) reported lower scores for their dog’s mental status, which in turn predicted poorer mental health. Cognitive biases commonly observed in individuals with insecure attachment types might influence perceptions of pet welfare whereby negative emotional states can predispose individuals to make more negative judgements about ambiguous social stimuli (in this case a pet’s behaviour and interactions as indicators of their mental status) thus increasing worry and concern, leading to poorer mental health [98]. Subjective biases in the perception of a pet’s quality of life, may have consequences for actual pet welfare [99], highlighting the importance of further investigation.

We were also interested in whether perceived pet behavioural problems explained the relationship between pet attachment and owner mental health. Our findings indicate that cat owners who scored high on attachment anxiety were more likely to report more cat behavioural problems and were more likely to report being bothered by these problems. Dog owners who scored high on attachment anxiety were also more likely to report more dog behavioural problems (total scores and all subscales except for attachment issues). This supports our previously proposed hypothesis that attachment insecurity could lead to negative perceptions regarding a pet’s behaviour and emotional state. Dog owners who scored high on attachment avoidance were more likely to report lower excitability and fewer attachment related issues, supporting our other theory that perhaps avoidantly attached individuals prefer and derive benefits from pets with particular characteristics. However, dog owners who scored high on attachment avoidance also reported more aggression and training difficulties, suggesting owner-pet attachment could influence dog behaviour [75]. This is supported by previous studies also finding an association between high avoidance in dog owners and owner-directed aggression; a theorised explanation being emotional distance, a lack of affection and availability from an owner could result in a lack of perceived secure base for the dog, evoking fear and thus aggression (see [76]). Higher reported dog behavioural problems did relate to worse mental health for dog owners in our study, with more dog fear and anxiety relating to increased owner depression and anxiety, and higher dog training difficulty relating to increased owner depression. Moreover, fear and anxiety in dogs partially mediated the relationship between anxious attachment and owner anxiety and depression. These findings support theory and past evidence that pet challenges could increase burden and influence owner mental health, suggesting that tailored support for pet behavioural issues could alleviate psychological distress [24, 32, 70, 97]. It is important to note however, that these findings were not replicated for cat owners despite past research demonstrating a link between cat behavioural problems and owner wellbeing [100]. It is also important to note again that we have focused on perceptions and self-reports of pet behavioural issues which may contain biases, and so these may not reflect accurate depictions of a pet’s behaviour. Future research could examine how to utilise more accurate assessments of a pet’s welfare and behaviour to gain a full picture of the possible impact on owner mental health, and what support is needed.

Finally, we explored whether perceived human-pet compatibility explained the relationship between pet attachment and owner mental health. Our findings revealed an association between high scores on attachment insecurity (both anxious and avoidant) and lower perceived total compatibility among both dog and cat owners. Specifically, dog owners who scored high on attachment anxiety reported lower compatibility across all domains, while avoidantly attached dog owners scored lower on all compatibility domains except for physical compatibility. Similarly, cat owners who scored high on anxious attachment scored lower on all compatibility domains except for physical and ‘other’, whereas avoidantly attached cat owners reported lower compatibility specifically in the affection and closeness domains. These findings indicate that insecurity within human-pet attachments could influence feelings of owner compatibility and thereby perceptions of the human-pet relationship. This supports our previously proposed theory that insecure pet attachment may lead to more negative misattributions of a pet’s behaviour, leading to feelings of emotional disconnection, unmet expectations and needs from the relationship, and relationship dissatisfaction. For example, insecurity within attachment relationships can lead to more negative expectations about a pet’s availability and responsiveness, as well as mistrust regarding their intentions [41, 42]. In relation to mental health, only the affection compatibility domain seemed to be important for dog owners, yet in reverse to our predictions, with those reporting higher compatibility also displaying higher anxiety scores. Affection compatibility also partially positively mediated the relationship between anxious attachment and dog owner anxiety, and fully negatively mediated the relationship between avoidant attachment and dog owner anxiety. Perhaps those with anxious attachments are more likely to seek out physical proximity and affection from their dogs, and may perceive such efforts as not being reciprocated, heightening feelings of rejection, which increases owner anxiety, whereas those with avoidant attachments do not have the same desire for physical closeness and affection from a pet [41, 42]. Placing high value on physical closeness and affection from a pet may indicate a lack of social support from wider human relationships, which can be a risk factor for poorer mental health [101], yet few studies have simultaneously accounted for the quality of human–human and human–pet relations when considering human wellbeing [102]. Further research is needed to disentangle the complex relationships between pet attachment orientations, owner mental health, and other relationship quality measures including compatibility, with human social support as a potential mediating variable.

A critical drawback of prior investigations into the mental health implications of pet attachment lies in the insufficient definition of attachment within human-animal relationships. Many studies fail to delineate attachment according to psychological attachment theory or employ measures that reliably evaluate attachment orientations. Instead, studies have focused on pet ownership alone, or the human-animal bond, overlooking the nuanced dimensions of attachment as outlined in psychological literature [29, 103–105]. A notable strength of our study lies in the utilization of a pet attachment measure grounded in established psychological theory, that is close to theoretical predications regarding attachment orientations, with pet relationships displaying the same structure of relationships as seen in human-human attachments [28]. The measure is derived from a validated and standardized instrument, aligning with the two-dimensional model of attachment observed within human-human relationships, enhancing the reliability of our findings (ECR-R) [79], (RQ) [80]. We found important findings relating to these attachment orientations, with attachment insecurity being a potential risk factor for poorer owner mental health, but also an influencing factor in perceptions of dog welfare and behaviour and perceived relationship compatibility. Future investigations should continue to delve into these relationship indicators, particularly in younger populations whereby reliable attachment measures are not currently available and development work is needed.

A further strength of our study lies in the exploration between dog and cat owners, revealing notable distinctions. However, a factor that we did not consider, is pet age, and duration of pet ownership. Attachment to a pet may develop overtime, and the benefits of such attachment for owner wellbeing may be more pronounced at certain time-points. For instance, the challenges and heightened stress associated with caring for puppies and kittens may delay the onset of wellbeing benefits until the pet has settled and a bond has formed [23]. Conversely, older pets may also present unique challenges, including concerns about pet health and anticipated grief [106].

A limitation of our investigation is that we did not consider social support from, or attachment orientations to, participant’s human relationships. As previously mentioned, social support and secure attachments to others, or lack thereof, could explain the impacts of pets for mental health, whereby those with high levels of social support and secure human attachments may have their needs saturated and thus have less need to seek support and emotional closeness from their pets [107]. Investigating socio-demographics differences was also out of the scope of the current investigation yet could yield noteworthy findings in future work. For example, those facing socioeconomic hardship can face increased pet burden and report more pet challenges, yet may not have the financial means to seek support, and may be reluctant to seek support due to stigma, exacerbating an already increased risk for mental health concerns [97, 108]. Also important is examining personal characteristics and identity. For example, 35% of our emerging adult sample identified as being LGBTQI+, and research has demonstrated that not only do such individuals still face increased risk of psychological distress due to prejudice, stigma, and discrimination, but that pets, and particularly the human-pet bond, can play an outsize role in buffering against some of these hardships, increasing resilience and decreasing stress (see [109]). Moreover, gender identity [15, 110] and owner personality characteristics [31, 47], could also play a role in human-pet relationships and so could also be considered in future work. Finally, other relationship concepts should be considered that have been overlooked until recently within the human-pet domain which could impact upon the human-pet relationship and owner mental health, including self-expansion, perceived pet responsiveness, and perceived pet insensitivity [29].

## Conclusion

This study reveals several nuanced findings about the complex relationship between pet ownership, owner-pet attachment, and owner mental health, indicating notable distinctions between dog and cat owners. The study also underscores the need to consider important owner-pet factors, such as owner-pet compatibility, pet welfare, and pet behavioural problems, to fully understand the impact of the human-pet relationship on the wellbeing of both owners and pets. Interventions that focus on promoting positive human-pet bonds, addressing negative cognitive biases, and mitigating pet welfare and behavioural issues, are likely to improve owner mental health. Additionally, managing pre-existing expectations around needs from a pet relationship, and considering compatibility when acquiring a pet, may also be crucial to improving the human-pet relationship, and subsequently, improving the wellbeing of both owners and their pets.

## Data Availability

The datasets presented in this article are not readily available because of participant privacy and ethical considerations. Requests to access the datasets should be directed to.

https://osf.io/s5ejy/

## Acknowledgements

A sincere thank you to all of the young adults who took their time to participate in this research.

## Author contributions

Conceptualization: RH

Data curation: RH, CR

Formal analysis: RH

Funding acquisition: RH

Investigation: RH, CR

Methodology: RH, CR

Project administration: RH, CR

Supervision: RH

Validation: RH

Visualization: RH

Writing – original draft: RH, AE

Writing – review & editing: AE, CR

